# Electrochemical Biosensing Platform based on Dual Detection of α-Synuclein and Quinolinic Acid as Neurological Diseases Biomarkers: Probe-less Point-of-Care Diagnostics of Early-Stage Parkinson’s Disease

**DOI:** 10.1101/2025.05.15.25327680

**Authors:** Bodapati Asritha, Athmakuri Tharak, S. Venkata Mohan

## Abstract

Neurological disorders like Parkinson disease (PD) present significant diagnostic challenges due to the lack of cost-effective and reliable biomarkers. This study aimed to develop an advanced electrochemical biosensing platform for detecting PD-associated biomarkers, alpha-synuclein and quinolinic acid, using multi-walled carbon nanotube (MWCNT)-modified screen-printed carbon electrodes (SPCEs). The objective was to create a probe-less, environmentally sustainable electrochemical sensor with high sensitivity and reproducibility for early PD diagnosis. MWCNTs were dispersed in a 1% sodium dodecyl sulfate solution and electrochemically deposited onto SPCEs, with scanning electron microscopy confirming a uniform, mesh-like nanostructure. Cyclic voltammetry (CV) and differential pulse voltammetry (DPV) were employed for biomarker detection in synthetic human serum. Quinolinic acid displayed a distinct oxidation peak at 0 V (12.54 microA), with DPV showing a linear response across 0.01 mM to 100 mM. Alpha-synuclein detection, enhanced by copper, revealed oxidative peaks at -0.26 V and -0.07 V, and reductive peaks at -0.52 V and -0.32 V, with DPV demonstrating concentration-dependent responses. The developed electrochemical sensors enable rapid, point-of-care (POC) testing for PD, requiring minimal sample volumes and offering portability for use in resource-limited settings. These sensors are crucial for early PD detection, facilitating timely intervention and monitoring disease progression. In conclusion, this probe-less electrochemical platform provides a cost-effective, sustainable tool for PD diagnostics, with significant potential to improve clinical outcomes through accessible and efficient POC applications.

## 1. Introduction

Neurological disorders, including Parkinson’s disease (PD), Alzheimer’s disease (AD), and multiple sclerosis (MS), constitute a major global health concern, collectively impacting millions of individuals worldwide. These conditions impose significant clinical and socioeconomic burdens and present complex challenges for healthcare systems due to their progressive nature, multifactorial etiology, and limited therapeutic options. (Negahdary et al., 2025; Carneiro et al., 2023; Khatami et al., 2024). These disorders are typified by progressive neuronal degeneration and functional impairment, often leading to debilitating motor, cognitive, and behavioural impairments. The complexity of their pathogenesis, coupled with the lack of definitive diagnostic tools, Emphasizes the pressing need for robust biomarkers to support early detection, monitor disease progression, and direct therapeutic strategies. Biomarkers, characterized as quantifiable indicators of biological states or conditions, are pivotal in elucidating the molecular mechanisms underlying neurological disorders and enhancing clinical outcomes. (Hassan et al., 2019; Parihar et al., 2024; Bagree et al., 2024). Among the promising biomarkers, alpha-synuclein and quinolinic acid have garnered significant attention due to their association with neurodegenerative processes, particularly in PD and related disorders.

Alpha-synuclein, a presynaptic protein, is a hallmark of PD pathology, primarily due to its aggregation into insoluble fibrils that form Lewy bodies, the pathological hallmark of the disease (Massey et al., 2023; Parihar et al., 2024). Its misfolding and aggregation are implicated in neuronal toxicity and synaptic dysfunction, driving disease progression. Quinolinic acid, a neurotoxic metabolite of the kynurenine pathway, is another critical biomarker, elevated in conditions such as PD, AD, and Huntington’s disease (İnce et al., 2024; da Silva et al., 2024; Kumar et al., 2022). Its excitotoxic properties contribute to neuronal damage by overstimulating N-methyl-D-aspartate (NMDA) receptors, leading to oxidative stress and inflammation (Kruszka et al., 2025; Malaiya et al., 2022). The accurate detection of these biomarkers in biological fluids, such as cerebrospinal fluid (CSF) or serum, is essential for early diagnosis and monitoring therapeutic efficacy. However, traditional detection methods, such as enzyme-linked immunosorbent assays (ELISA), mass spectrometry, and immunohistochemistry, face significant limitations, including high costs, lengthy processing times, large sample volumes, and the need for sophisticated instrumentation. These drawbacks hinder their widespread clinical application, particularly in resource-limited settings.

Recent advancements in electrochemical biosensing have emerged as a promising alternative for biomarker detection, offering high sensitivity, selectivity, and rapid response times. Electrochemical techniques, including cyclic voltammetry (CV) and differential pulse voltammetry (DPV), leverage the redox properties of analytes to generate measurable electrical signals, enabling the detection of biomarkers at low concentrations. These methods are particularly advantageous for detecting alpha-synuclein and quinolinic acid, as they can be tailored to recognize specific molecular interactions, such as the binding of alpha-synuclein with copper ions, which exacerbates protein aggregation in PD. Copper, a transition metal, plays a pivotal role in modulating alpha-synuclein pathology by binding to its N-terminal region, promoting misfolding, and accelerating the formation of toxic oligomers. This interaction has been extensively studied, with research highlighting its relevance to PD pathogenesis (Wang et al., 2010; Lee et al., 2008; Rose et al., 2011; Castillo-Gonzalez et al., 2017).

The development of electrochemical sensors for neurological biomarkers has also been driven by the need for point-of-care (POC) diagnostics, which prioritize low-cost, portable, and user-friendly platforms (Bae et al., 2025; Sajeevan et al., 2025; Narváez et al., 2025). Traditional electrochemical approaches often rely on complex electrode modifications, such as the incorporation of molecular probes, aptamers, or antibodies, to enhance specificity. While effective, these modifications increase costs, require specialized expertise, and may introduce environmental concerns due to the use of hazardous chemicals. Moreover, probe-based systems can suffer from stability issues, limiting their long-term applicability.

To address these challenges, present study introduces a novel electrochemical approach for the detection of alpha-synuclein and quinolinic acid, leveraging a probe-less electrode modification strategy that emphasizes simplicity, cost-effectiveness, and environmental sustainability. By utilizing screen-printed carbon electrodes modified with MWCNTs dispersed in SDS, this method eliminates the need for expensive probes while maintaining high sensitivity and reproducibility. The use of synthetic human serum as a testing matrix enhances the clinical relevance of the approach, requiring minimal sample volumes and enabling rapid analysis. The incorporation of copper in alpha-synuclein detection further enhances specificity by exploiting its role in protein aggregation, offering insights into PD pathophysiology. This study’s novelty lies in its streamlined electrode modification process, which combines low-cost materials, environmentally friendly techniques, and robust electrochemical performance, positioning it as a promising tool for the early diagnosis and monitoring of neurological disorders.

## 2. Experimental Methodology

### 2.1 Chemicals & reagents

The chemicals, copper (II) chloride, quinolinic acid (CAS number: 89-00-9), α synuclein human recombinant E. coli (575001), sodium dodecyl sulfate, multiwalled carbon nanotubes, and, Phosphate Buffered Saline (pH 7.5) along with those required for the preparation of synthetic human serum were all procured from Sigma Aldrich Co. Private Limited.

### 2.2 Protein integrity

The integrity of the alpha-synuclein protein was assessed using SDS-PAGE electrophoresis, following the protocol detailed by Tharak et al., 2025 and Bhattacharjee et al. (2023). Briefly, protein samples were denatured by heating at 95°C for 5 minutes in Laemmli sample buffer containing 2% (w/v) SDS, 10% (v/v) glycerol, 62.5 mM Tris-HCl (pH 6.8), 5% (v/v) β-mercaptoethanol, and 0.01% (w/v) bromophenol blue. Denatured samples were separated on a 12% polyacrylamide gel at 120 V for 90 minutes using a running buffer of 25 mM Tris, 192 mM glycine, and 0.1% (w/v) SDS (pH 8.3). Post-electrophoresis, the gel was stained with 0.1% (w/v) Coomassie Brilliant Blue R-250 in 40% methanol and 10% acetic acid for 1 hour. Protein bands were visualized after destaining in a solution of 10% methanol and 10% acetic acid. The molecular weight and integrity of alpha-synuclein were confirmed by comparing its migration pattern to a pre-stained protein ladder, ensuring the protein’s stability and purity for subsequent experiments.

### 2.3 MWCN Electro-polymerization

Screen-printed carbon electrodes (SPCEs) were procured from Zensor R&D and subjected to a systematic modification process to enhance their analytical performance for electrochemical sensing applications. To ensure homogeneous dispersion of multi-walled carbon nanotubes (MWCNTs), a 1% (w/v) sodium dodecyl sulfate (SDS) solution was formulated in deionized water. The surfactant properties of SDS were utilized to reduce surface tension, ensuring a stable and homogeneous MWCNT suspension. Specifically, 1 mg of MWCNTs was dispersed in 1 mL of the 1% SDS solution (1 mg/mL), and the mixture was sonicated at a frequency of 53 kHz and a temperature of 60°C for 30 minutes. These sonication parameters were optimized to effectively disentangle MWCNT bundles, promoting their uniform distribution in the suspension. Prior to modification, the SPCEs were thoroughly cleaned to ensure a contaminant-free surface. The electrodes were washed with a 2:1 (v/v) ethanol-deionized water solution, followed by rinsing with deionized water and air-drying at ambient temperature. This cleaning step was critical to remove any organic residues and impurities, providing a pristine substrate for MWCNT deposition. Subsequently, 10 µL of the MWCNT-SDS dispersion was drop-cast onto the working electrode surface of the SPCE. Electrochemical deposition of MWCNTs was then performed using cyclic voltammetry, with 15 consecutive potential cycles applied between -1.0 V and 1.0 V (vs. Ag/AgCl) at a scan rate of 50 mV/s in a three-electrode configuration, carbon served as both the working and counter electrodes, with Ag functioning as the reference electrode.

The controlled electrochemical deposition process facilitated the formation of a stable and uniform MWCNT layer on the SPCE surface, as confirmed by the consistent voltammetric response across cycles. This method ensured strong adhesion of the MWCNT layer, which is essential for reproducible electrochemical sensing and biosensing applications. The use of SDS as a surfactant provided a cost-effective and environmentally sustainable approach for MWCNT dispersion, outperforming alternative surfactants in terms of dispersion stability and uniformity. Furthermore, the sonication step enhanced dispersion efficiency, contributing to a well-defined MWCNT layer that improved the sensitivity and reliability of the modified electrodes. Compared to conventional modification techniques such as physical adsorption or drop-casting, the electrochemical deposition method employed in this study offered superior control over the MWCNT layer thickness and morphology. Additionally, the probe-less modification strategy presented economic and environmental benefits, eliminating the need for costly probe materials and reducing waste generation.

### 2.4 SPCE Morphology

The evaluation of electrode surface morphology was carried out using a scanning electron microscope (SEM) to assess the effectiveness of depositing multi-walled carbon nanotubes (MWCNTs). Comparative analyses were undertaken between the unmodified electrode and the electrode with MWCNT deposition to identify structural differences. The magnification levels utilized in this study spanned from 25x to 20,000x, providing a comprehensive view of the surface alterations induced by MWCNT deposition. This detailed examination facilitated a deeper understanding of the nanomaterial’s influence on electrode morphology and its potential implications for electrochemical applications.

### 2.5 Electrochemical analysis

The experimental methodology employed a Metrohm Autolab Potentiostat model, specifically the PGSTAT-204, in combination with the NOVA 2.1 software suite to conduct comprehensive data analysis (Tharak et al., 2024; Madamanchi et al., 2025; Anitha et al., 2004). The electrochemical analysis was conducted using a working volume of 40 µl, carefully chosen to adequately cover the electrode’s entire surface area. This volume selection process involved considerations for optimal electrode coverage, enhancing the reliability of the results. Differential pulse voltammetry (DPV) and cyclic voltammetry (CV) techniques were employed to investigate biomarker identifications and their corresponding redox potentials. The scan rate for cyclic voltammetry was set at 20 mV/s and the potential window ranged from -1 V to 1 V, allowing for comprehensive electrochemical assessments within this specified range. During the differential pulse voltammetry analysis, a series of voltage pulses with a step size of 0.005 V were applied to generate voltammograms. This finer step size selection aimed to produce sharper and more discernible peaks, enhancing the precision of biomarker identification. The potential range for DPV spanned from -1.2 V to 1.2 V, providing a wider scope for redox potential analysis. To ensure the reliability and reproducibility of the electrode’s performance, data averaging from three cycles was implemented.

## 3. Results and discussion

### 3.1 synuclein integrity and 3-D stricture

The SDS-PAGE electrophoresis analysis successfully confirmed the integrity of the alpha-synuclein protein. Following the separation of denatured protein samples on a 12% polyacrylamide gel, distinct protein bands were visualized through Coomassie Brilliant Blue R-250 staining. Alpha-synuclein was identified at its expected molecular weight of 10 kDa by comparing its migration pattern to a pre-stained protein ladder, indicating that the protein remained intact and undegraded (Fig. 1a). This verification of purity and integrity was critical prior to proceeding with further experiments. Additionally, the structural folding of alpha-synuclein was evaluated using the SWISS-MODEL homology modeling tool with the following sequences: MDVFMKGLSK AKEGVVAAAE KTKQGVAEAA GKTKEGVLYV GSKTKEGVVH GVATVAEKTK EQVTNVGGAV VTGVTAVAQK TVEGAGSIAA ATGFVKKDQL GKNEEGAPQE GILEDMPVDP DNEAYEMPSE EGYQDYEPEA. The predicted three-dimensional structure revealed a predominantly disordered conformation with regions of transient helical propensity, consistent with the intrinsically disordered nature of alpha-synuclein reported in the literature (Uversky et al., 2001).

**Figure 1.**
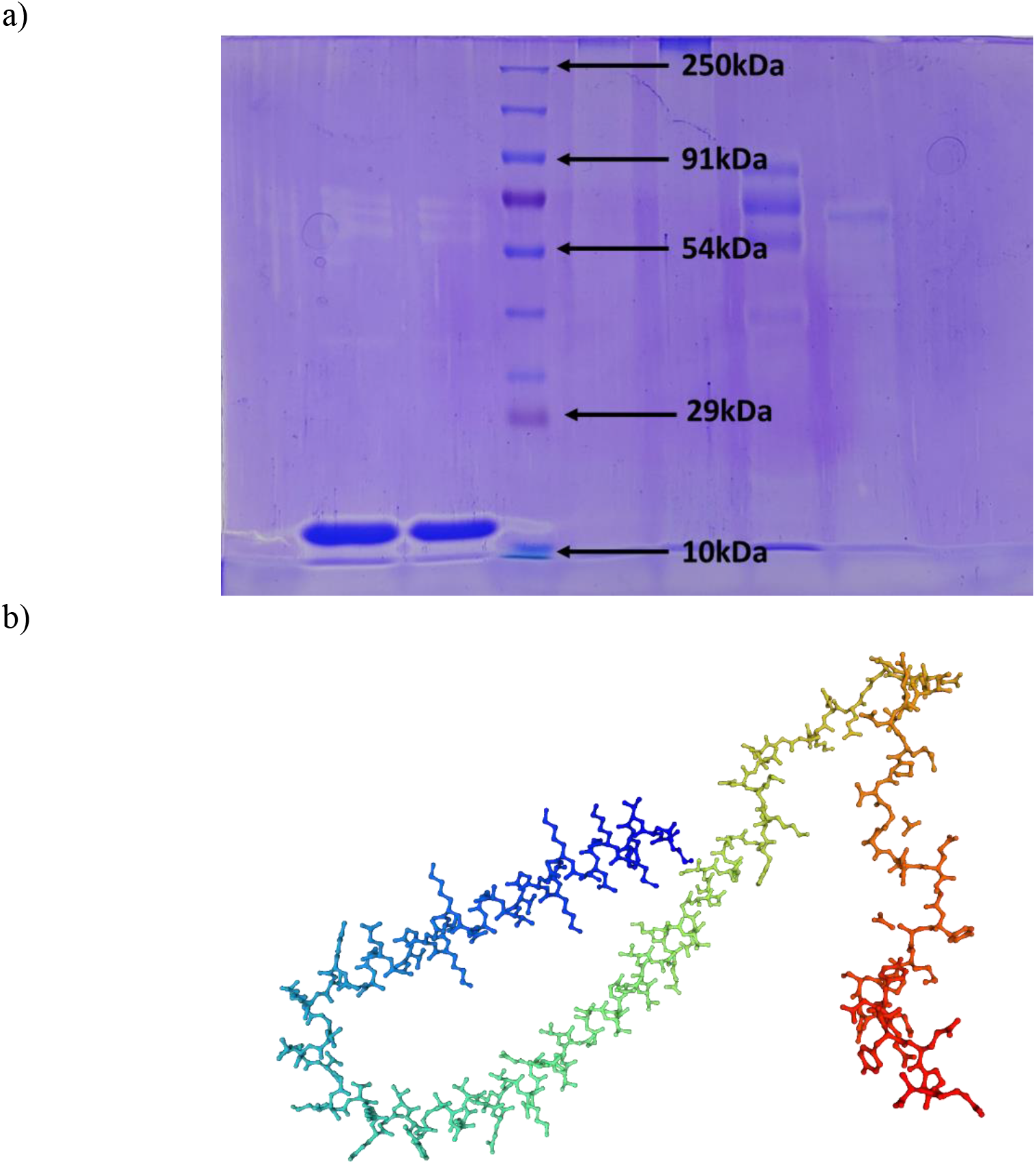
Characterization of α-synuclein protein. (a) SDS-PAGE analysis of α-synuclein protein integrity on a 12% polyacrylamide resolving gel, stained with Coomassie Brilliant Blue R-250, confirming the protein’s molecular weight at 10 kDa. (b) Three-dimensional structural model of α-synuclein generated using SWISS-MODEL, illustrating a predominantly disordered conformation with regions of transient helical propensity.

### 3.2 Impact of MWCNT on the Morphological Characteristics of SPCE

SEM analysis revealed precise structural features that confirmed the successful deposition of multi-walled carbon nanotubes (MWCNTs) on the screen-printed carbon electrode (SPCE),evidenced by comparing the non-deposited SPCEs morphology (Fig. 2a-b). The SEM images demonstrated a uniform and well-distributed MWCNT layer across the electrode surface, highlighting the effectiveness of the sonication process in achieving homogeneous dispersion. At a magnification of 25×, the modified electrode exhibited a mesh-like structure, providing evidence of the successful integration of MWCNTs into the electrode matrix (Fig. 2c). Further examination at higher magnifications offered deeper insights into the interaction between MWCNTs and the electrode substrate, revealing a cohesive layer that significantly enhanced the electrode’s surface area and conductivity. At magnifications of 5000× (Fig. 2d) and 10,000× (Fig. 2e), a clustered arrangement of MWCNTs was clearly observed, underscoring the robustness of the electrochemical deposition process. At 20,000× magnification (Fig. 2f), the characteristic tubular nanostructure of the MWCNTs became distinctly visible, providing detailed insights into the nanomaterial’s architecture at the microscale level. These findings validate the efficacy of the MWCNT deposition technique and elucidate the morphological attributes that contribute to the improved functionality and performance of the modified SPCE.

**Figure 2.**
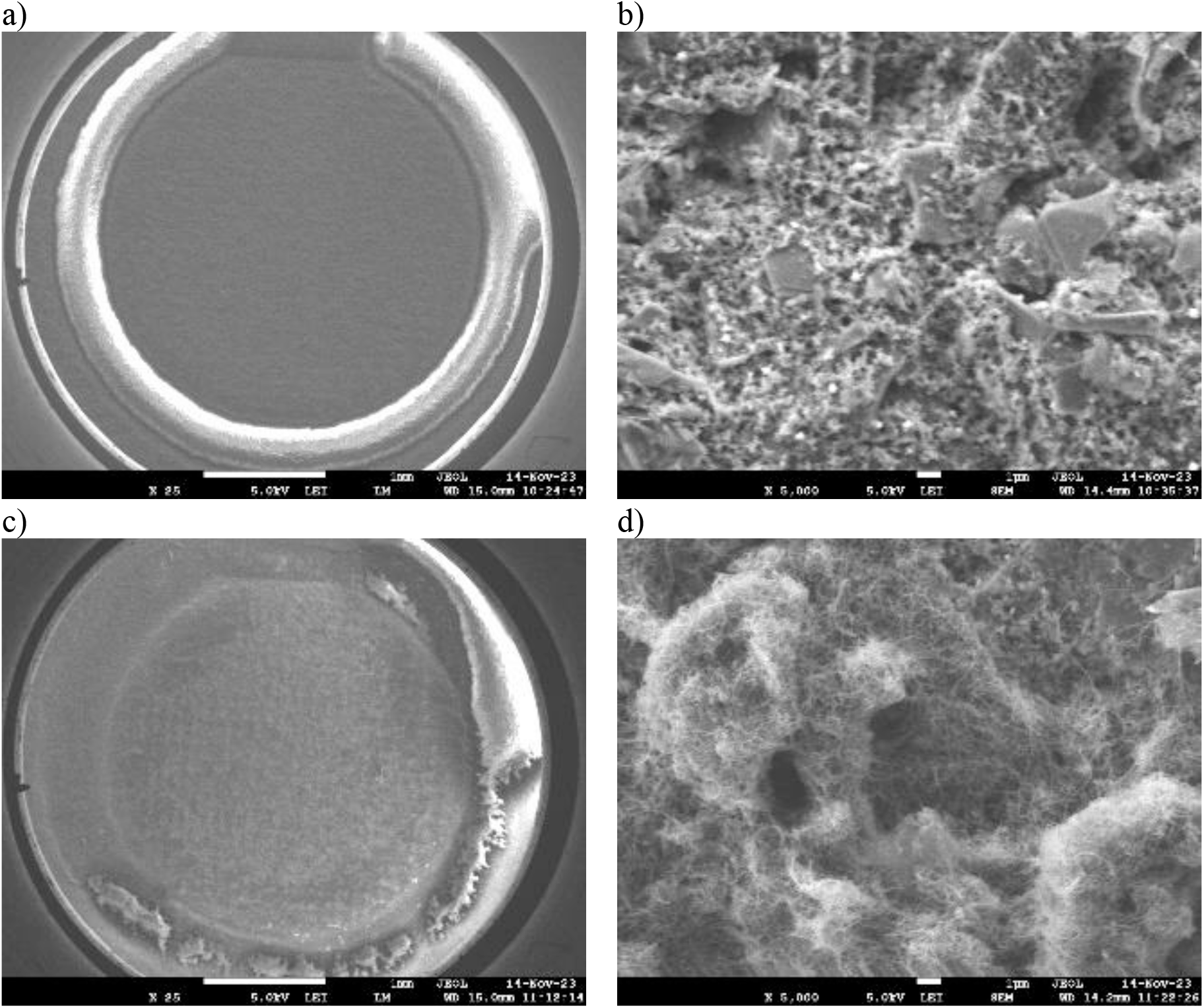
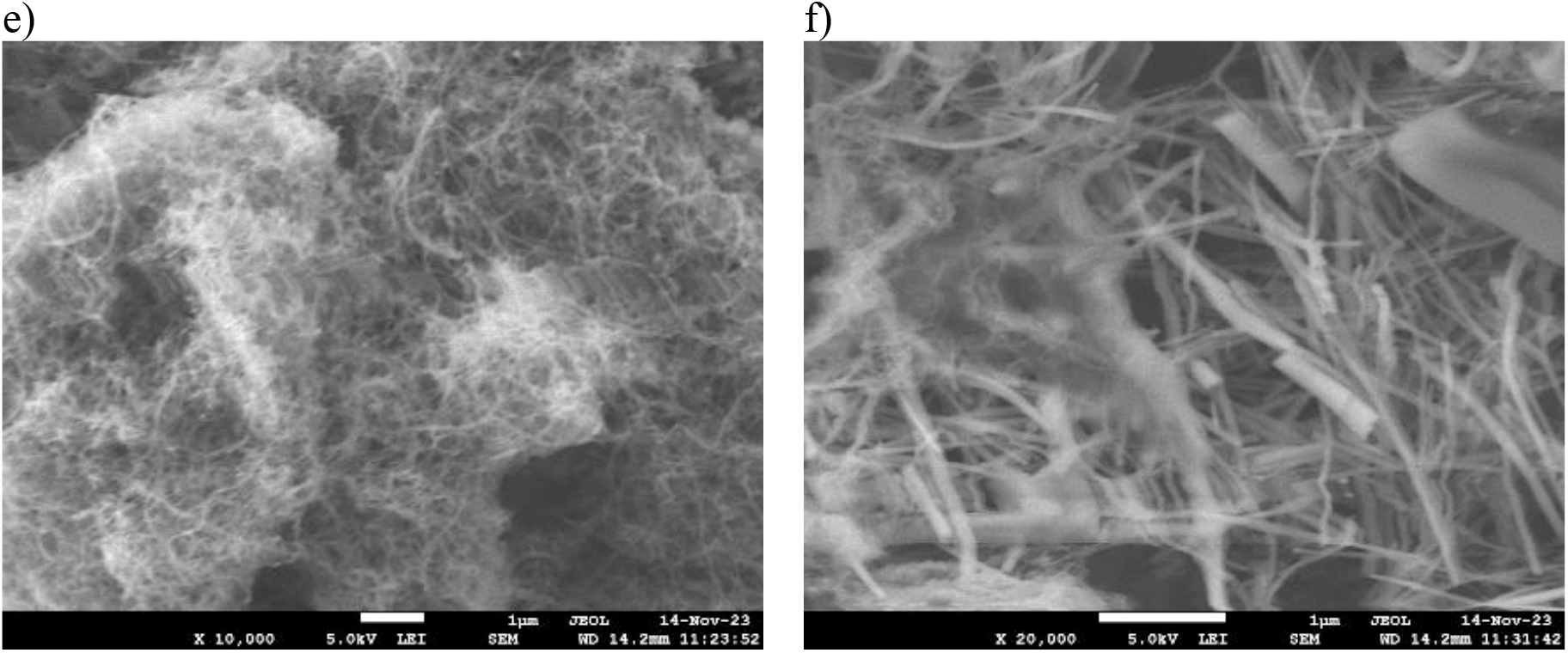
SEM images comparing bare and MWCNT-modified screen-printed carbon electrodes (SPCEs). (a) Bare electrode at 25× magnification, showing the pristine surface morphology. (b) Bare electrode at 5000× magnification, highlighting surface details. (c) MWCNT-modified electrode at 25× magnification, revealing a mesh-like structure indicative of successful MWCNT deposition. (d) MWCNT-modified electrode at 5000× magnification, displaying a clustered arrangement of MWCNTs. (e) MWCNT-modified electrode at 10,000× magnification, further illustrating the nanotube distribution. (f) MWCNT-modified electrode at 20,000× magnification, clearly depicting the characteristic tubular nanostructure of MWCNTs.

### 3.3 Electrochemical Detection and Quantification of Biomarker 1-Quinolinic acid

The uniform deposition of multi-walled carbon nanotubes (MWCNTs) on the working electrode of the screen-printed carbon electrode (SPCE) was verified prior to initiating the electrochemical identification of quinolinic acid. To establish a baseline, initial electrochemical analyses were conducted using a pristine solution of medical-grade water as the blank medium, which was free of solutes that could interfere with the detection process. Cyclic voltammetry of the blank solution revealed no discernible peaks, confirming its suitability as a control. Subsequently, quinolinic acid was introduced into the blank solution, and cyclic voltammetry was performed. The resulting voltammogram exhibited a distinct oxidation peak at 0.2 V, consistent with findings reported by Singh et al. (2017), confirming the presence of quinolinic acid. This peak is depicted in the cyclic voltammograms provided in Fig. S1 of the Supplementary Information.

To ensure the reliability of the cyclic voltammetry data, measurements were averaged over three cycles to minimize random fluctuations and enhance result robustness. Between each cycle, the electrode was thoroughly cleaned with a 2:1 (v/v) ethanol-water solution, followed by a blank solution run to eliminate potential deposition or contamination that could affect subsequent measurements. Following baseline establishment, a synthetic human serum solution was prepared according to the protocol by Basiaga et al. (2014) and stored at 4°C. Cyclic voltammetry of the synthetic human serum alone displayed a reduction peak at -0.2 V. Upon addition of quinolinic acid to the synthetic human serum, a distinct oxidation peak was observed at approximately 0 V, with a peak current of 12.54 µA. Additionally, the capacitance of the voltammogram decreased significantly following the addition of quinolinic acid (Fig. 3a). This electrochemical behavior is likely attributable to the redox properties of quinolinic acid and its interactions with components in the synthetic human serum. The observed oxidation peak at 0 V suggests the oxidation of quinolinic acid, potentially influenced by interactions with the electrode surface or serum components, while the decrease in capacitance may result from alterations in the solution’s dielectric properties induced by quinolinic acid.

**Figure 3:**
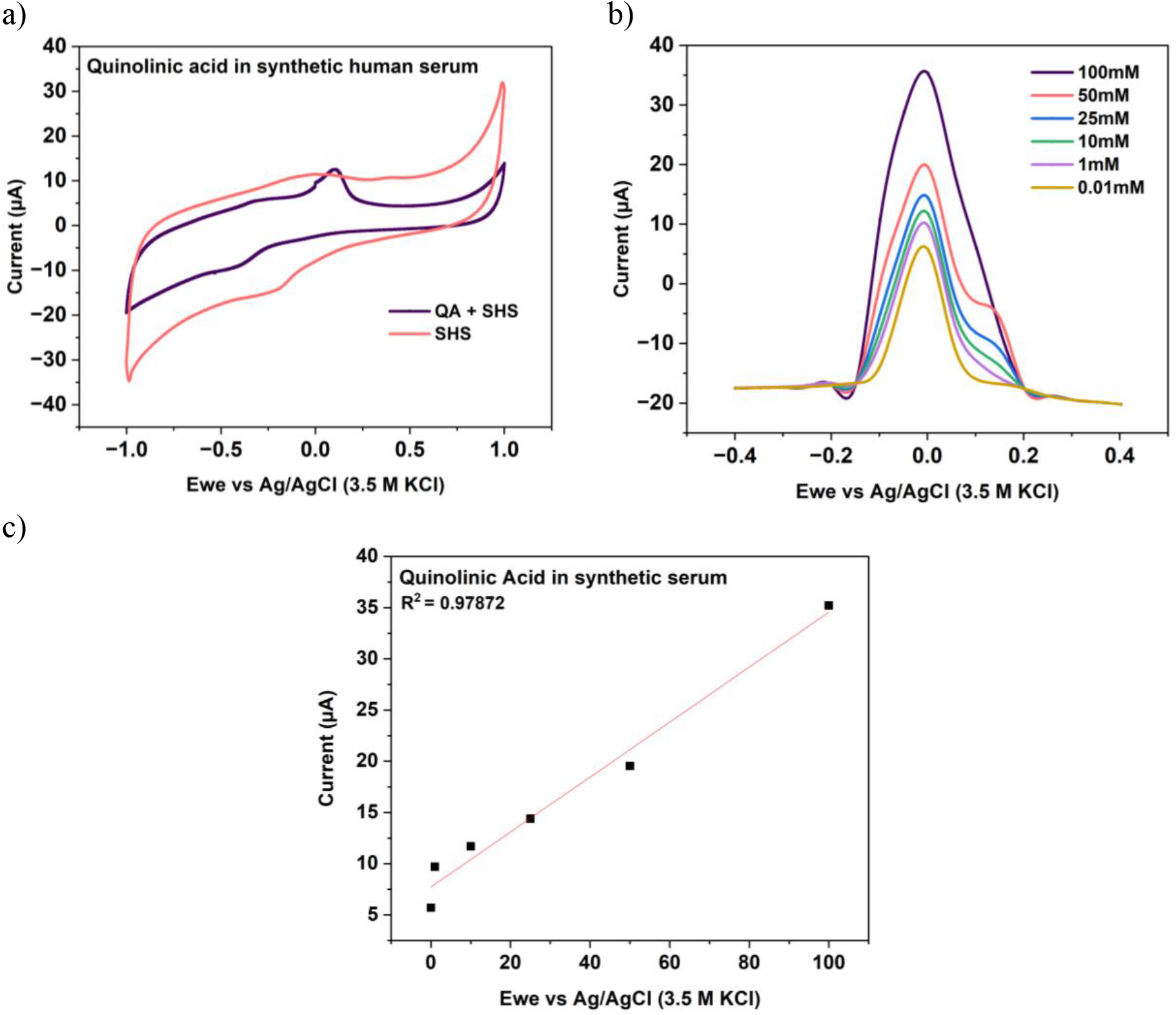
Electrochemical analysis of quinolinic acid in synthetic human serum. (a) Cyclic voltammograms comparing synthetic human serum alone (reduction peak at -0.2 V) with synthetic human serum containing quinolinic acid (oxidation peak at 0 V, peak current of 12.54 µA). (b) Differential pulse voltammograms of quinolinic acid in synthetic human serum at concentrations of 0.01 mM to 100 mM, with peak currents increasing from 5.7 µA to 35.22 µA. (c) Calibration curve of quinolinic acid in synthetic human serum, plotting peak current versus concentration, yielding an R^2^ value of 0.98.

To further investigate the relationship between quinolinic acid concentration and electrochemical response, differential pulse voltammetry (DPV) was employed. Quinolinic acid in the concentrations of 0.01 mM to 100 mM were tested in the synthetic human serum. At the lowest concentration of 0.01 mM, the peak current was recorded at 5.7 µA, while at the highest concentration of 100 mM, the peak current increased to 35.22 µA (Fig. 3b). This ten thousand-fold increase in concentration corresponded to an approximately six-fold increase in peak current, demonstrating the sensitivity of the DPV method for detecting quinolinic acid across a wide concentration range. A calibration curve was constructed using the peak currents, yielding a high correlation coefficient (R^2^ = 0.98) (Fig. 3c), indicating a strong linear relationship between quinolinic acid concentration and peak current. The consistency and reproducibility of the peak currents across multiple trials, facilitated by the rigorous electrode cleaning protocol between cycles, further validate the robustness and accuracy of the experimental methodology for quantifying quinolinic acid in a synthetic human serum matrix.

### 3.4 Electrochemical Detection of Biomarker-2-Alpha-synuclein and Copper Interactions

The next biomarker investigated was α-synuclein, a protein critically concerned in Parkinson’s disease (PD) pathology. Initial experiments focused on the electrochemical detection of α-synuclein alone; however, the inclusion of copper ions significantly enhanced the detection process due to copper’s role in promoting α-synuclein aggregation (Castillo-Gonzalez et al., 2017). Previous studies have established that copper ions specifically bind to the N-terminus of α-synuclein, inducing conformational changes that lead to protein misfolding and aggregation (Stefanis, 2012). This interaction results in the formation of toxic oligomers and fibrils, which are precursors to Lewy bodies, contributing to neuronal dysfunction and degeneration in PD (Rose et al., 2011; Srinivasan et al., 2021). Additionally, post-translational modifications such as phosphorylation of α-synuclein have been shown to increase its binding affinity for copper, further accelerating the aggregation process (Castillo-Gonzalez et al., 2017). Understanding the complex interplay between copper and α-synuclein aggregation is essential for elucidating PD pathogenesis and developing targeted therapeutic strategies.

CV was employed to characterize the electrochemical properties of α-synuclein and copper individually. Pure α-synuclein exhibited a reduction peak at -0.58 V with a peak current of -39.27 µA (Fig. 4a). However, these measurements were challenging due to the protein’s inherent instability, necessitating low-temperature conditions to ensure accurate and reproducible results. This instability highlighted the importance of incorporating copper into the detection system. A copper solution was prepared by dissolving copper ions in phosphate-buffered saline (PBS) at pH 7.0, and its electrochemical behavior was assessed. The cyclic voltammogram of the copper solution displayed a distinct oxidation peak at -0.17 V with a peak current of 2.09 µA and a reduction peak at -0.38 V with a peak current of -0.49 µA (Fig. 4b). These electrochemical signatures of copper emphasize its critical role in modulating α-synuclein aggregation and its relevance to PD pathophysiology.

**Figure 4:**
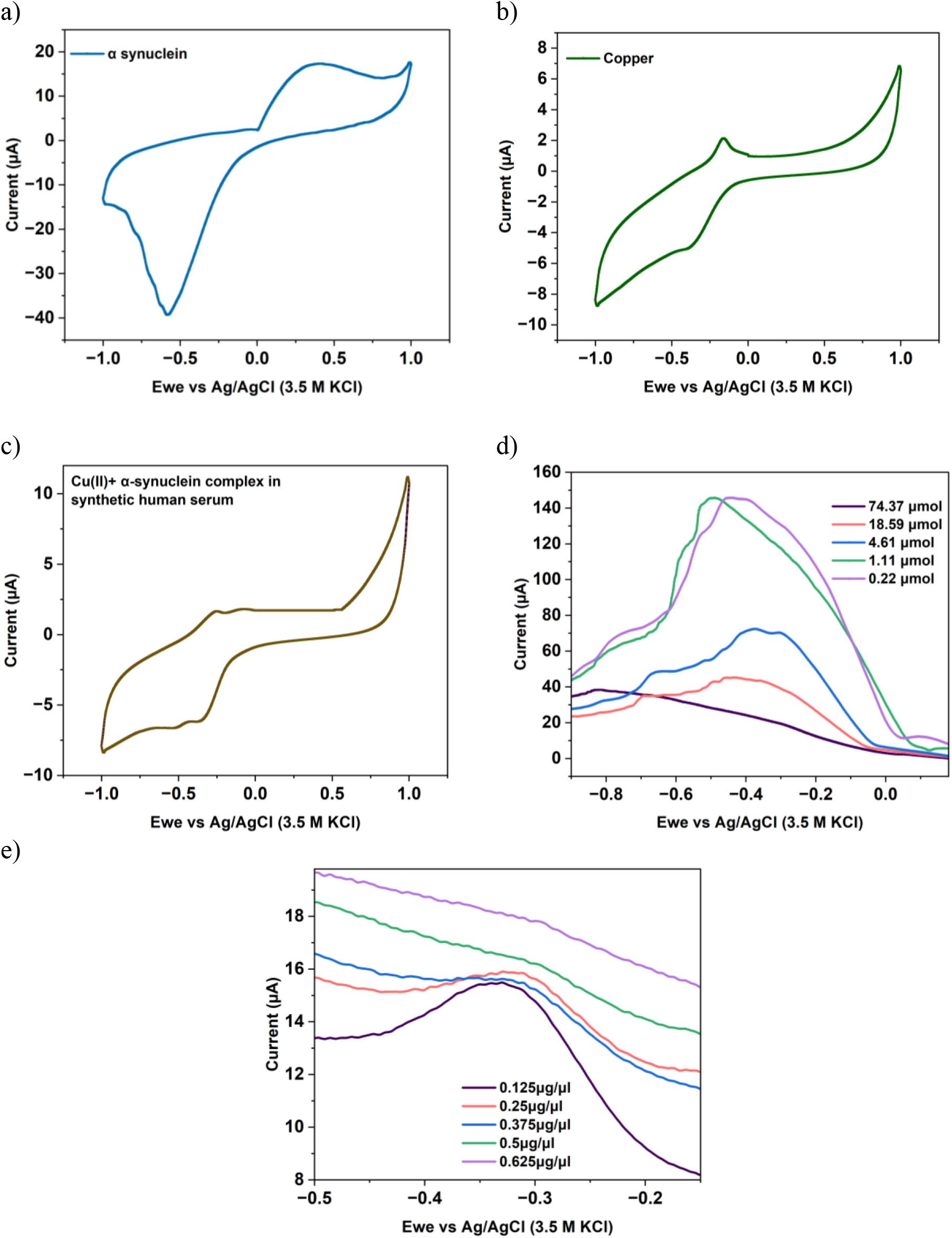
Electrochemical characterization of α-synuclein and copper interactions. (a) Cyclic voltammogram of pure α-synuclein in phosphate-buffered saline (PBS), exhibiting a reduction peak at -0.58 V. (b) CV of copper in PBS, depicting oxidation peak at -0.17 V and a reduction peak at -0.38 V. (c) CV of a mixture of copper and α-synuclein in synthetic human serum at a 1:1 (v/v) ratio, displaying oxidative peaks at -0.26 V and -0.07 V, and reductive peaks at -0.52 V and -0.32 V. (d) Differential pulse voltammogram in synthetic human serum with a constant α-synuclein concentration, where copper concentrations were varied from 10 µg/mL to 0.03 µg/mL, showing voltage peaks between -0.6 V and -0.3 V. (e) Differential pulse voltammogram in synthetic human serum with a constant copper concentration, where α-synuclein concentrations were varied from 0.125 µg/µL to 0.625 µg/µL, displaying voltage peaks between -0.5 V and -0.2 V.

A solution containing α-synuclein and copper was mixed with synthetic human serum at a 1:1 (v/v) ratio, and cyclic voltammetry was performed to evaluate their combined electrochemical behavior. The resulting voltammogram revealed two oxidative peaks at -0.26 V and -0.07 V, with corresponding peak currents of 1.63 µA and 1.80 µA, respectively. Additionally, two reductive peaks were noticed at -0.52 V and -0.32 V, with peak currents of -6.63 µA and -6.18 µA, respectively (Fig. 4c). These electrochemical signatures suggest the partial formation of an α-synuclein-copper complex, which likely contributes to the observed redox behavior in the synthetic human serum matrix. To further investigate the interaction dynamics, experiments were conducted by varying copper concentrations while keeping the α-synuclein concentration constant in the synthetic human serum. Copper concentrations were gradually decreased from 10 µg/mL (74.37 µmol) to 0.03 µg/mL (0.22 µmol). Electrochemical analysis revealed voltage peaks within the range of -0.6 V to -0.3 V (Fig. 4d), with a linear regression analysis yielding an R^2^ value of 0.9211 (Fig. S2A). Notably, the α-synuclein peak current increased as copper concentration decreased, indicating a potential shift in the equilibrium of the protein-copper complex formation. Conversely, when α-synuclein concentrations were incrementally increased from 0.125 µg/µL to 0.625 µg/µL while maintaining a constant copper concentration, electrochemical analyses showed voltage peaks between -0.5 V and -0.2 V (Fig. 4e). The corresponding R^2^ value was 0.7022 (Fig. S2B), and the protein-copper complex peaks exhibited a steady increase with rising α-synuclein concentrations. These findings suggest a reversible interaction between copper and α-synuclein, consistent with previous studies on their redox reactions and binding properties (Wang et al., 2010). Copper ions are known to incorporate into α-synuclein amyloids, primarily interacting with residues in the N-terminal region (Lee et al., 2008), although the extent of this interaction may vary depending on specific experimental conditions and concentrations of copper and α-synuclein.

## 4. Conclusions

This study successfully demonstrated the efficacy of a novel electrochemical biosensing platform for the detection of quinolinic acid and α-synuclein, key biomarkers associated with Parkinson’s disease (PD), using multi-walled carbon nanotube (MWCNT)-modified screen-printed carbon electrodes (SPCEs). The probe-less electrode modification strategy with MWCNTs significantly enhanced electrode surface area and conductivity, improving the sensitivity of the biosensing platform. Electrochemical analyses, employing CV and DPV, enabled sensitive and selective detection of quinolinic acid in synthetic human serum, with a strong linear relationship between concentration and peak current, affirming the method’s quantitative reliability. The incorporation of copper in α-synuclein detection highlighted its role in protein aggregation, with distinct electrochemical signatures indicating the formation of a protein-copper complex. The reversible interaction between α-synuclein and copper, influenced by their respective concentrations, provided valuable insights into PD pathophysiology. This cost-effective and environmentally sustainable approach, eliminating the need for complex probe-based systems, offers a promising tool for the early diagnosis and monitoring of neurological disorders. The methodology’s simplicity and robustness position it as a viable candidate for point-of-care diagnostics in clinical settings.

## Data Availability

All data produced in the present work are contained in the manuscript

## Acknowledgments

The authors would like to acknowledge the Director, CSIR-IICT (Manuscript No. IICT/Pubs./2025/178.) for the support.

